# Prevalence of psychiatric disorders among children and adolescents in Nigeria since 2010: A systematic review and meta-analysis

**DOI:** 10.1101/2025.02.22.25322718

**Authors:** Samuel Busayo Ogunlade, Ajibola Ibraheem Abioye, Moshood Abiodun Kuyebi, Jolaade Musa, Aminu Kende Abubakar, Mohammed Nakodi Yisa, Sarah Oreoluwa Olukorode, Oluwafemi Temitayo Oyadiran, Temitayo Rebecca Okusanya, Daniel Oluwafemi Olofin, Ebuwa Igho-Osagie, Moshood Olanrewaju Omotayo, Abiodun Adewuya

## Abstract

**Background:** Psychiatric disorders significantly impact pediatric populations globally, affecting individual development, education and societal integration. In Nigeria, where nearly half of the population is under the age of 15, there remains a substantial gap in our understanding of the burden of these disorders within this demographic. This systematic review was conducted to quantify the prevalence of psychiatric disorders among Nigerian children and adolescents in clinical and community settings.

**Methods:** We identified and examined original research articles available in PUBMED/Medline, EMBASE, and AJOL from January 2010 to August 2024, and selected studies that evaluated the prevalence of psychiatric disorders among children and adolescents (<18 y) in Nigeria. We pooled the prevalence estimates across different study settings using random effects models We assessed the risk of bias using the NIH Quality Assessment Tool and the certainty of the evidence using the GRADE framework.

**Results:** The meta-analysis included data from 27 studies with a total population of 47,451 participants. The best evidence of the prevalence was 12% for major depression (95% CI: 5.3, 25.0; I^2^ = 99.5%; seven studies; 14,534 children and adolescents; very low certainty), and 9.2% for generalized anxiety disorder (95% CI: 4.5, 17.9; I^2^ = 95.9%; five studies among 2,395 individuals; very low certainty). Meta-regression indicated that the prevalence of major depression was related to age among children (*p-*heterogeneity = 0.0004). The pooled prevalence of ADHD among primary school pupils was 2.8% (95% CI: 2.0, 3.8%; I² = 0%; two studies; 1,374 individuals). Overall, the pooled prevalence estimates of psychiatric disorders among Nigerian pediatric populations indicated considerable heterogeneity across most studies (I² > 75%). The most common psychiatric disorders across the different settings were major depression among children and adolescents attending primary care (prevalence = 7.6%; 95% CI: 1.0; 40.5; three studies; 1,278 individuals), separation anxiety disorder among primary school children (prevalence = 14%; 95% CI: 9.5, 19.6; one study; 200 pupils), and behavioral disorders among secondary school students (prevalence = 15.1%; 95% CI: 14.4, 15.8; one study; 9,450 adolescents).

**Conclusion:** The prevalence of psychiatric disorders among Nigerian children and adolescents is substantial, underscoring the critical need for enhanced mental health services.

## Introduction

Mental health disorders among pediatric populations have emerged as a significant public health concern globally, with an increasing recognition of their prevalence and impact on children’s development, academic performance, and overall well-being ^1^. Studies have shown that children with untreated mental health disorders are twice as likely to suffer developmental delays ^2^, have an 18.5% higher dropout rate from school ^3^, and have a 2-3 times higher risk of entering adulthood with chronic health conditions like epilepsy, diabetes and asthma ^4^. They are also less likely to be employed ^5^ and more likely to encounter legal issues as adults ^6^. According to the World Health Organization (WHO), approximately 10-20% of children and adolescents experience mental disorders, with half of all mental illnesses beginning by the age of 14 ^7^. For instance, anxiety disorders are among the most common mental health conditions in children and adolescents, with global prevalence rates ranging from 6 – 20%^8,9^. Depression affects about 2 – 8% of children and adolescents worldwide and is a leading cause of disability in this age group^10^.

Despite the substantial burden these disorders place on children, their families, and the healthcare system ^11–13^, there is a paucity of comprehensive data on the prevalence and trends of mental health disorders among Nigerian children. According to the National Bureau of Statistics, children under the age of 15 constitute approximately 44% of the Nigerian population^14^. Yet, there is a severe shortage of mental health professionals who focus on pediatric care, inadequate funding for mental health services, and pervasive stigma associated with mental health disorders, which collectively hinder the effective delivery of care ^15,16^. Previous studies on pediatric mental health in Nigeria have often been limited by small sample sizes, regional focus, and variability in diagnostic criteria, leading to fragmented and inconsistent data. Unfortunately, large-scale country-wide epidemiological surveys may be very expensive and complicated to conduct at the necessary level of rigor, quality control, and validity across the country. A systematic and comprehensive review of well-conducted regional studies of mental health disorders among Nigerian children and adolescents may be easier and cheaper to guide the design of clinical and policy interventions.

To the best of our knowledge, there are no previously reported systematic reviews or meta-analyses on the prevalence of childhood or adolescent mental disorders in Nigeria. This systematic review and meta-analysis aims to bridge the existing knowledge gap by synthesizing available data on the prevalence of mental health disorders in Nigerian pediatric populations. By critically evaluating and integrating findings from various studies, we provide a clearer understanding of the scope and scale of these disorders and identify potential risk factors.

## Methods

To the extent possible, this study adhered to the Preferred Reporting Items for Systematic Review and Meta-Analysis (PRISMA) checklist^17^ and the Joanna Briggs Institute (JBI) methodology for systematic reviews on prevalence ^18^. The protocol was developed and submitted to PROSPERO^19^ (Registration number: CRD 42024559090)

### Information sources

A comprehensive systematic search of PUBMED/Medline (U.S. National Library of Medicine), EMBASE (Elsevier), and African Journals Online (AJOL) was undertaken. Our primary focus was on the current prevalence of mental health disorders among the general and clinical populations of children and adolescents in Nigeria. Therefore, publications from January 2010 until August 2024 were examined.

#### Search strategy

The search strategy employed a combination of MeSH (Medical Subject Headings) terms, Emtree terms, and indexing topics for the PubMed/Medline and Embase databases, respectively. Relevant text and keywords were also utilized to enhance the search comprehensiveness. A hand-search of the references of selected articles and related systematic reviews was conducted to identify pertinent studies not captured through database searches.

#### Study Selection

Studies of mental disorders among children and adolescents in Nigeria were included – whether they were diagnosed by a physician or using a validated diagnostic tool. To the extent possible, we followed the classification and definition of disorders in the Diagnostic and Statistical Manual of Mental Disorders, Fifth edition (DSM-V) ^20^. Included studies had to have been designed to be representative of the general community or specific demographic or clinical subgroups with well-defined inclusion characteristics e.g., children living with HIV. Participants in the primary studies had to be <18 years old or students at a primary or secondary school. Systematic reviews, meta-analyses, or other review papers were excluded. Studies focusing on animals, cell lines, and case-control studies were also not considered. However, studies with cross-sectional design or surveys, and baseline data from cohort studies or trials were included. Studies addressing past or lifetime history of mental illness were excluded due to concerns about recall.

Study selection was conducted in two stages: an initial title/abstract review to exclude non-qualifying studies followed by a full article review of non-excluded studies to determine eligibility. Study selection was performed independently by two investigators, with any disagreements resolved through discussion with a third reviewer.

#### Data Extraction

Data extraction was conducted independently by assigned investigators using a standardized Excel form. Extracted data included diagnostic methodologies, participation sampling of the study population, details of the specific mental health disorders, study location, setting (e.g., schools, healthcare facilities, communities), demographic characteristics, study period, study name, prevalence percentages, number of cases, and the number of non-cases.

#### Risk of bias in individual studies

Risk of bias was evaluated using the National Institutes of Health (NIH) Quality Assessment Tool for Observational Cohort and Cross-Sectional Studies ^21^. This tool assessed whether the population was clearly defined, non-response was minimal, and a consistent sampling approach was applied. It also evaluated whether the sample was representative, the sample size was appropriately determined, and the mental health disorder was clearly defined.

### Data Synthesis

Quantitative synthesis was conducted for studies providing data on the prevalence of mental health disorders and associated risk factors. The primary summary measure was the pooled prevalence rate of mental health disorders, using the prevalence proportion as the effect measure in a random-effects meta-analysis model to accommodate study variability. Heterogeneity was assessed using the I² statistic, categorized as minimal (0-40%), moderate (>40-60%), substantial (60-80%), or considerable (>80%) ^22^.

Sensitivity and subgroup analyses were conducted to test the robustness of the review findings and explore sources of heterogeneity based on patient demographics, study settings, and geographic locations. Influence analysis compared the pooled estimate and I² statistic on leaving out studies sequentially. Baujat plots visualized the relationship of each study’s influence on the summary proportion to its contribution to the overall heterogeneity represented by the Cochran *Q*-statistic ^23^. In cases where quantitative synthesis was not feasible, a narrative synthesis summarized the key findings, focusing on prevalence rates and risk factors for mental health disorders, and identifying research gaps and future directions.

Publication bias was not assessed as the conventional funnel plots and Egger’s statistical tests used for this purpose do not apply to prevalence meta-analyses ^23^.

#### Confidence in Cumulative Evidence

The quality of the evidence for each prevalence measure was systematically assessed using a modification of the GRADE approach ^24^. The certainty of evidence was downgraded if included studies were not representative (risk of bias), if the pooled estimate was imprecise (imprecision), if point estimates and confidence intervals from each study or group varied substantially from others (inconsistency), or if the assessed population or outcome assessment differs from the population or outcome assessment of interest (indirectness). For imprecision, the following evidence thresholds based on a rule of thumb were used: 50%, 30%, 10%, 5%, 1%, 0.10 and 0%, as they change clinical and public health perceptions of how prevalent or rare the condition is. A summary grade of moderate or high certainty indicated confidence that the true prevalence was accurately estimated or close to the estimated prevalence, while a low or very low certainty suggested that the true prevalence was probably meaningfully different from the estimated prevalence ^25^.

#### Sensitivity analysis

In sensitivity analysis, we included studies conducted since 2006, instead of 2010 as was done in the main analysis. The most recent nationally representative survey was published in 2006^26^.

#### Data management

Database records were exported to a Zotero library for managing search results and removing duplicates, Rayyan for study selection^27^, a custom Microsoft Excel spreadsheet for data extraction, and Google Sheets for collaborative record-keeping.

## Results

27 studies examined the prevalence of pediatric mental illness in Nigeria since 2010 and were included in this review (**Figure 1).** These studies were conducted among 47,451 people and across Nigeria’s 37 states/administrative units. The highest number of studies were from Oyo (n=8), Lagos (n=7), and Enugu (n=6), excluding the multi-center studies conducted in multiple states.

**Figure 1.**
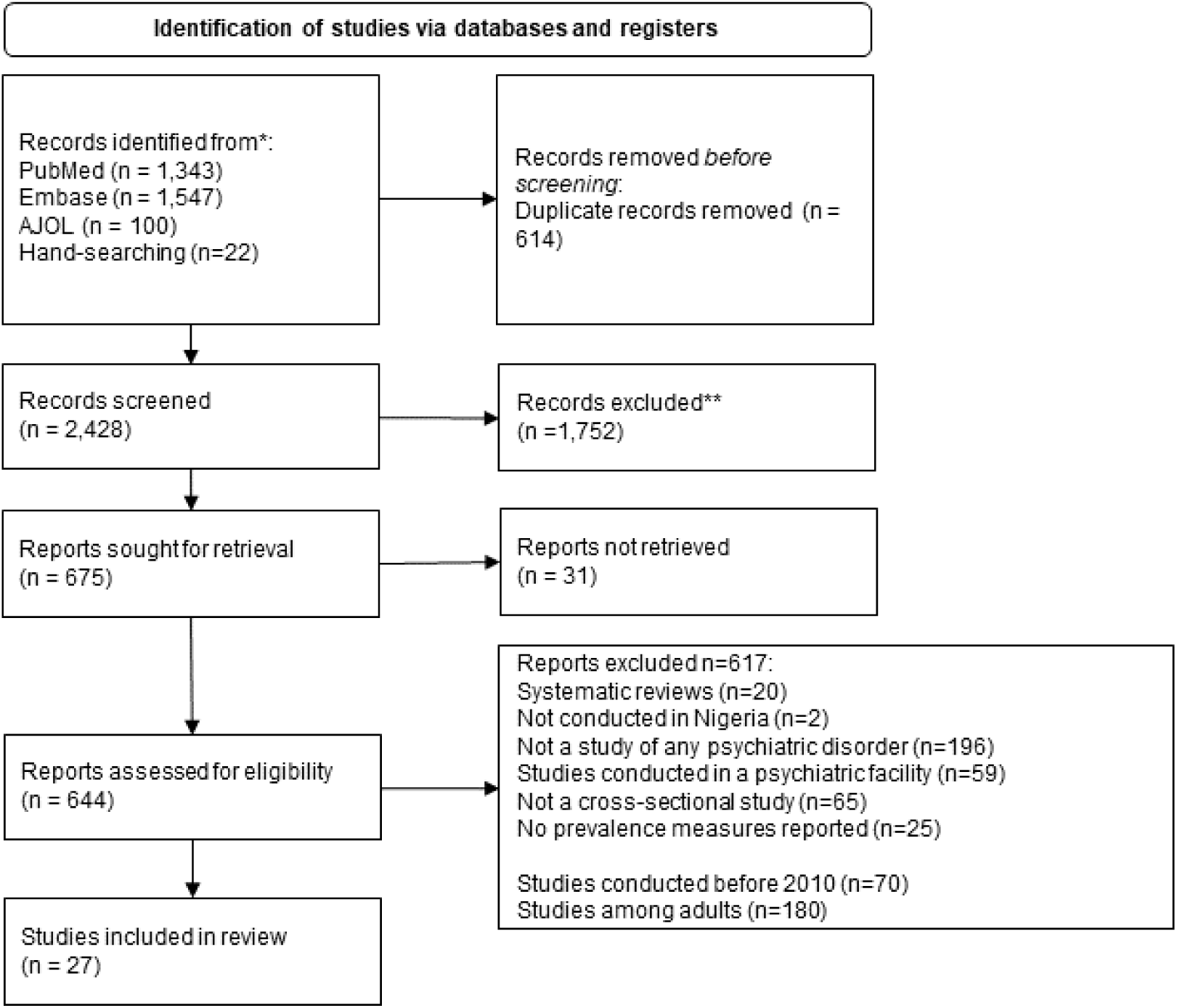
Flowchart of study selection

### Depressive disorders

17 studies assessed major depression, the clinically significant form of depression (**Table 1).** These studies were conducted in primary/elementary school^28^, secondary school ^29–35^, primary care settings ^36–38^, teen mothers ^39^, and among children with HIV ^40–42^, and epilepsy^43^.

**Table 1:** Depressive disorders among children and adolescents in Nigeria.

| Outcome | Population studied | Prevalence<br>(95% CI) | Sample size<br>(Number of<br>studies) | States | Certainty of<br>evidence | Comments<sup>1-4</sup> |
| --- | --- | --- | --- | --- | --- | --- |
| Major<br>depression | Primary school <sup>28</sup> | 0.4%<br>(0.1, 1.0) | 1,174<br>(1 study) | Akwa Ibom | ⊕⊕⊕⊕<br>High |  |
|  | Secondary school <sup>29-35</sup> | 17.1%<br>(10.9, 25.8) | 13,360<br>(7 studies) | Ebonyi, Enugu,<br>Lagos, Oyo, SW<br>states | ⊕⊕⊕○<br>Moderate | Inconsistency |
|  | Primary care <sup>36-38</sup> | 7.6%<br>(1.0, 40.5) | 1,278<br>(3 studies) | Kwara, Ogun,<br>Oyo | ⊕○○○<br>Very low | Inconsistency and<br>extreme imprecision |
|  | People with: |  |  |  |  |  |
|  | HIV <sup>40-42</sup> | 15.1%<br>(12.1, 18.7) | 456<br>(3 studies) | Cross River,<br>Lagos, Oyo,<br>Osun | ⊕⊕⊕⊕<br>High |  |
|  | Sickle cell disease <sup>44</sup> | 8.3%<br>(3.4, 16.4) | 84<br>(1 study) | Enugu | ⊕⊕○○<br>Low | Serious imprecision |
|  | Epilepsy <sup>43</sup> | 30.8%<br>(26.2, 35.7) | 380<br>(1 study) | Rivers | ⊕⊕⊕○<br>Moderate | Imprecision |

1. Inconsistency: Confidence intervals from a study or group varied substantially from others 2. Imprecision: Downgraded as 95%CI included a wide range with different clinical interpretations 3. Serious imprecision: Downgraded twice as 95%CI included a very wide range with substantially different clinical interpretations 4. Extreme imprecision: Downgraded three times as 95%CI included an extremely wide range with considerably different clinical interpretations

**Table 2:** Mood and anxiety disorders among children and adolescents in Nigeria.

| Outcome | Population studied | Prevalence<br>(95% CI) | Sample size<br>(Number of studies) | States | Certainty of<br>evidence | Comments<sup>1, 2</sup> |
| --- | --- | --- | --- | --- | --- | --- |
| Generalized anxiety disorder | Primary school <sup>45</sup> | 8.0%<br>(4.6, 12.7) | 200<br>(1 study) | Kaduna | ⊕⊕○○<br>Low | Serious imprecision |
|  | Almajiri school <sup>45</sup> | 16.0%<br>(11.3, 21.6) | 213<br>(1 study) | Kaduna | ⊕⊕⊕○<br>Moderate | Imprecision |
|  | Secondary school <sup>31,35,46</sup> | 9.5%<br>(3.9, 21.4) | 2,195<br>(3 studies) | Anambra,<br>Ebonyi, Osun | ⊕⊕○○<br>Low | Serious imprecision |
|  | Primary care <sup>38</sup> | 1.1%<br>(0.3, 2.9) | 350<br>(1 study) | Kwara | ⊕⊕○○<br>Low | Serious imprecision |
| Bipolar I disorder manic episode | Primary school <sup>45</sup> | 0.5%<br>(0.01, 2.6) | 200<br>(1 study) | Kaduna | ⊕⊕○○<br>Low | Serious imprecision |
|  | Almajiri school <sup>45</sup> | 1.0%<br>(0.1, 3.6) | 213<br>(1 study) | Kaduna | ⊕⊕○○<br>Low | Serious imprecision |
| Agoraphobia | Primary school <sup>45</sup> | 0.5%<br>(0.01, 2.6) | 200<br>(1 study) | Kaduna | ⊕⊕○○<br>Low | Serious imprecision |
|  | Almajiri school <sup>45</sup> | 1.0%<br>(0.1, 3.6) | 213<br>(1 study) | Kaduna | ⊕⊕○○<br>Low | Serious imprecision |
| Social phobia | Primary school <sup>45</sup> | 2.5%<br>(0.8, 5.7) | 200<br>(1 study) | Kaduna | ⊕⊕○○<br>Low | Serious imprecision |
|  | Almajiri school <sup>45</sup> | 0.9%<br>(0.1, 3.4) | 213<br>(1 study) | Kaduna | ⊕⊕○○<br>Low | Serious imprecision |
| Separation<br>anxiety | Primary school <sup>45</sup> | 14.0%<br>(9.5, 19.6) | 200<br>(1 study) | Kaduna | ⊕⊕⊕○<br>Moderate | Imprecision |
|  | Almajiri school <sup>45</sup> | 1.4%<br>(0.3, 4.1) | 213<br>(1 study) | Kaduna | ⊕⊕○○<br>Low | Serious imprecision |

1. Imprecision: Downgraded as 95%CI included a wide range with different clinical interpretations
2. Serious imprecision: Downgraded twice as 95%CI included a very wide range with substantially different clinical interpretations

The pooled prevalence of major depression among children and adolescents attending secondary school was approximately 17.1% (95% CI: 10.9%, 25.8%; I^2^ = 99%; **Figure 2A**) based on seven studies among 13,360 people^29–35^. **Supplementary Figure 1** shows the relationship of the influence of the individual studies on the summary proportion against their contribution to the overall heterogeneity. The pooled prevalence did not change by >20% on excluding any studies. The I^2^ did not also change meaningfully. The certainty of the evidence was moderate – downgraded for inconsistency. The included studies all utilized different diagnostic instruments, including the PHQ-9 ^30,35^. The pooled prevalence from two studies that used the PHQ-9 was 21.7% (95% CI: 20.0, 23.4).

**Figure 2.**
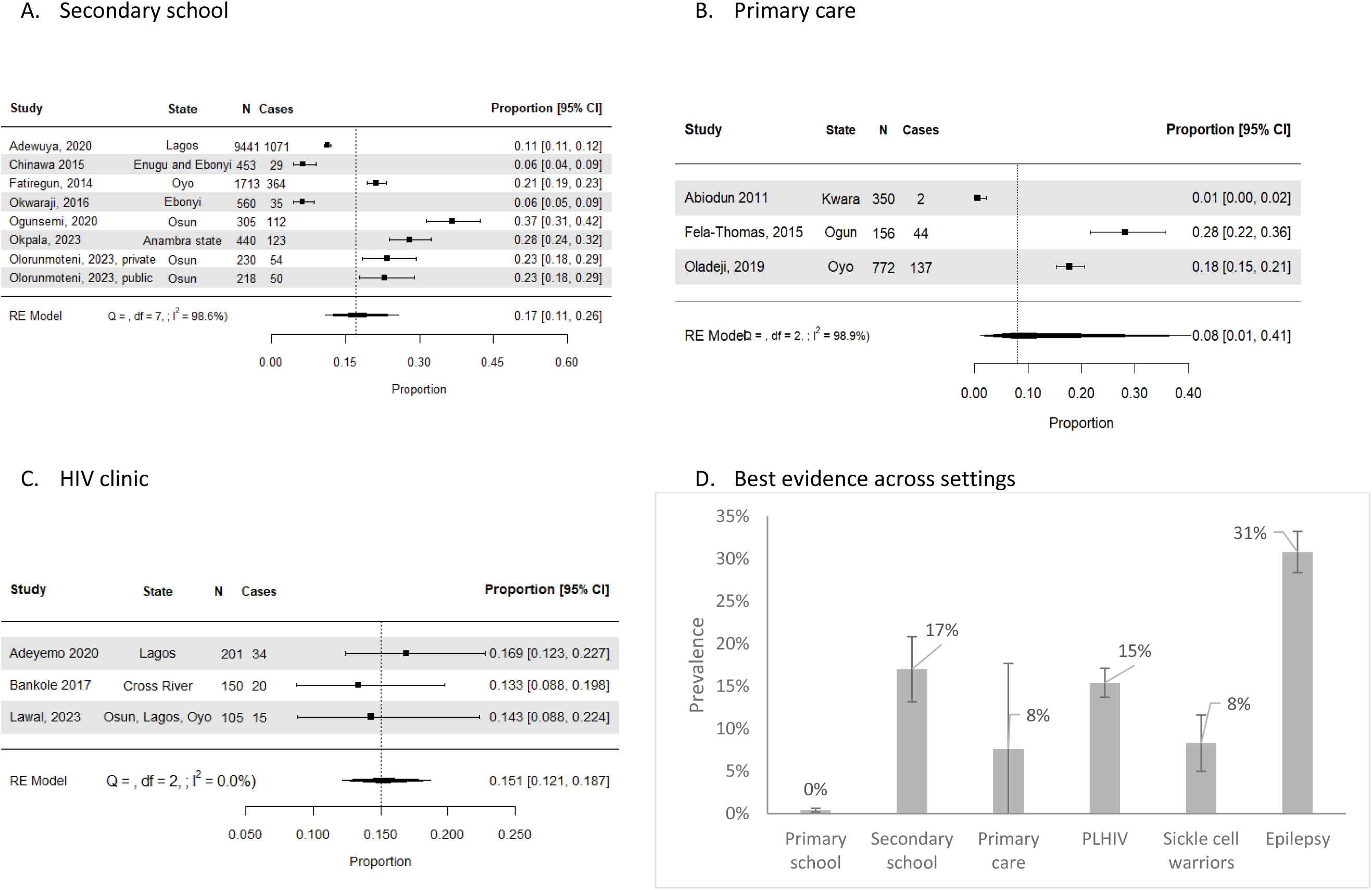
Major depression among young children and adolescents in secondary school in Nigeria

**Table 3:** Neurodevelopmental disorders.

| Outcome | Population studied | Prevalence (95% CI) | Sample size (Number of studies) | States | Certainty of evidence | Comment<sup>1,2</sup> |
| --- | --- | --- | --- | --- | --- | --- |
| Neurodevelopmental disorders |  |  |  |  |  |  |
| ADHD | Primary school<sup>28,45</sup> | 2.8%<br>(2.0, 3.8) | 1,374<br>(2 studies) | Akwa Ibom,<br>Kaduna | ⊕⊕⊕⊕<br>High |  |
|  | Almajiri school<sup>45</sup> | 0.5%<br>(0.01, 2.6) | 213<br>(1 study) | Kaduna | ⊕⊕○○<br>Low | Serious imprecision |
|  | Primary care<sup>38</sup> | 1.4%<br>(0.5, 3.3) | 350<br>(1 study) | Kwara | ⊕⊕⊕○<br>Moderate | Imprecision |
| Mental retardation | Primary care<sup>38</sup> | 0.6%<br>(0.1, 2.0) | 350<br>(1 study) | Kwara | ⊕⊕⊕○<br>Moderate | Imprecision |
| Autism spectrum disorders | Primary and secondary schools<sup>51</sup> | 2.9%<br>(1.8, 4.4) | 721<br>(1 study) | Enugu and<br>Ebonyi | ⊕⊕⊕⊕<br>High | Imprecision |

1. Imprecision: Downgraded as 95%CI included a wide range with different clinical interpretations
2. Serious imprecision: Downgraded twice as 95%CI included a very wide range with substantially different clinical interpretations

**Table 4:**
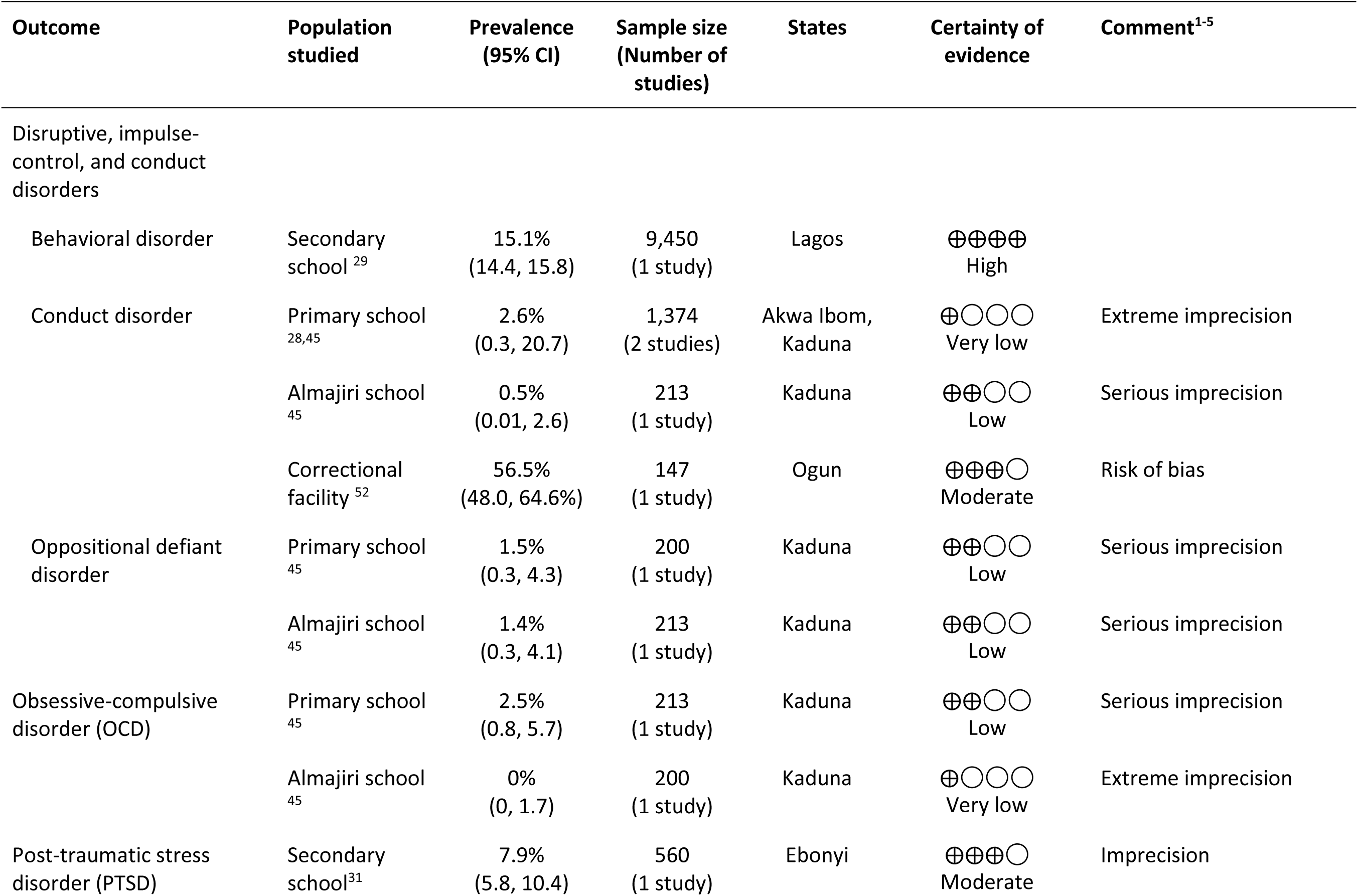
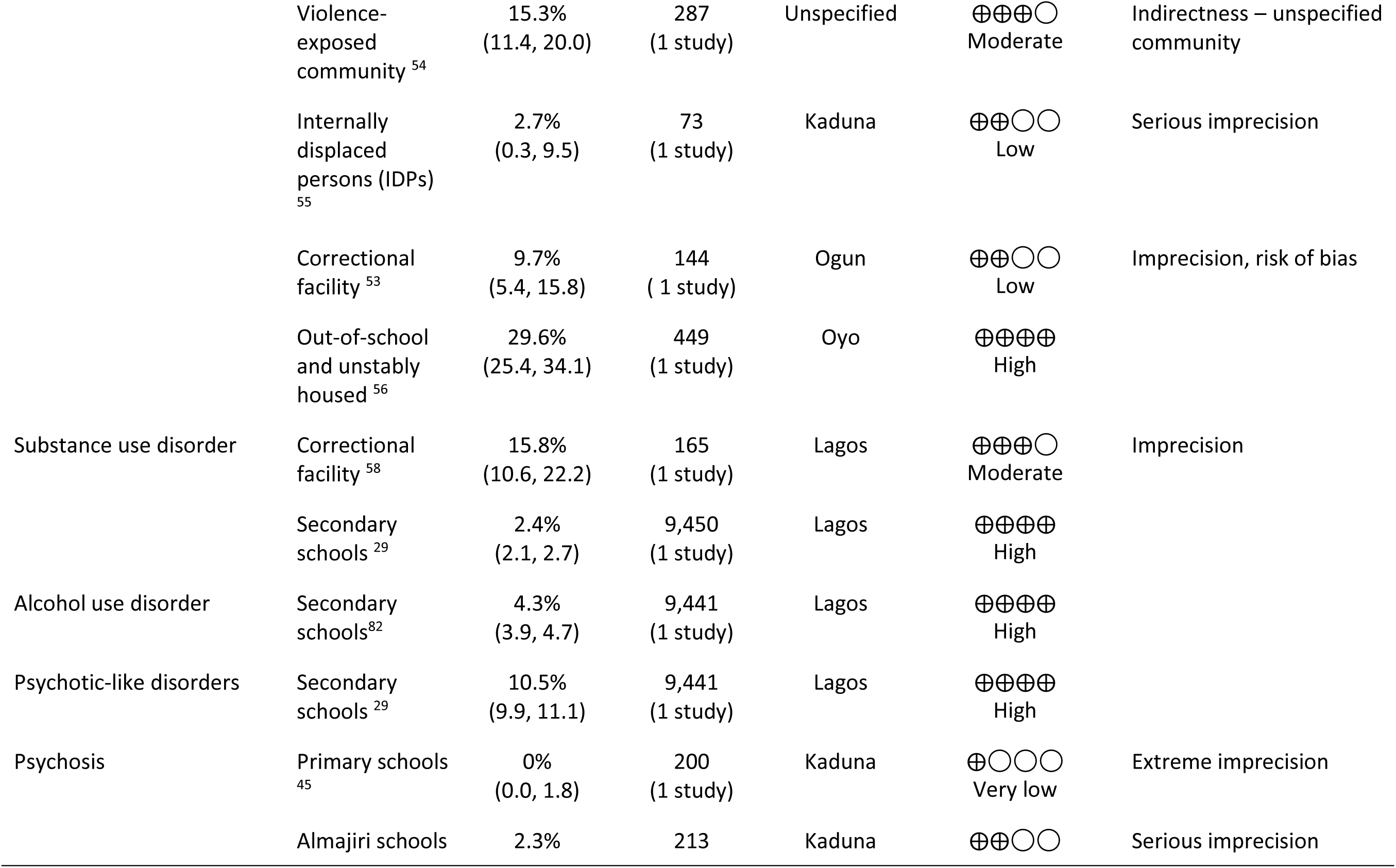
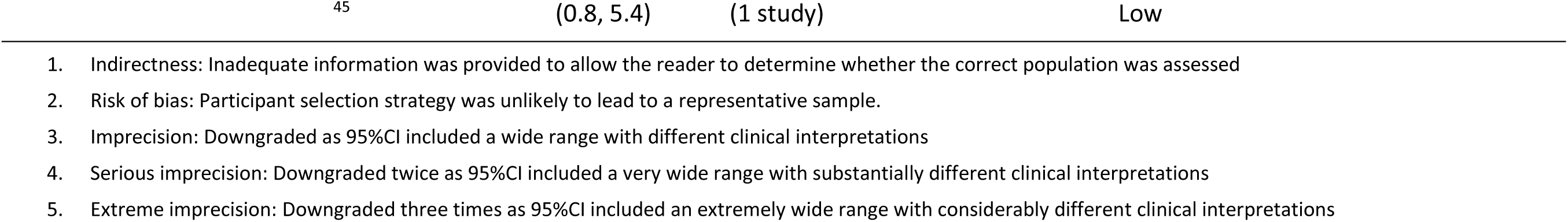
Other mental disorders.

The pooled prevalence of major depression among children and adolescents in primary care settings was approximately 7.6% (95% CI: 1.0%, 40.5%; I^2^ = 99%; **Figure 2B**) based on three studies among 1,278 people^36–38^. Influence analysis was not considered as there were too few studies. The certainty of the evidence was very low – downgraded for inconsistency and extreme imprecision.

The pooled prevalence of major depression among children and adolescents with HIV was approximately 15.1% (95% CI: 12.1, 18.7; I^2^ = 0%; **Figure 2C**) based on three studies among 456 people^40–42^. The certainty of evidence was high.

Low certainty evidence from single studies also indicates that the prevalence of major depression was 0.4% (95% CI: 0.1, 1.0) among 1,174 primary school pupils^28^, 8.3% among 84 children with sickle cell disease(36), and 30.8% among 380 children and adolescents with epilepsy^43^. **Figure 2D** presents the prevalence of major depression based on the best available evidence across the settings studied.

Using the data from settings most representative of the general population of children and adolescents – primary and secondary schools, in the absence of community studies, the pooled prevalence of major depression was 12.0% (95% CI: 5.3, 25.0; I^2^ = 99.5%; seven studies among 14,534 individuals). The certainty of the evidence was very low – downgraded for serious imprecision and inconsistency. Meta-regression indicates that the prevalence of major depression in Nigeria is significantly related to age among children and adolescents in Nigeria (*p-*heterogeneity = 0.0004; **Figure 3)**. Major depression is typically first recognized around age 9, and the prevalence increases considerably as adolescence begins especially from age 12.

**Figure 3.**
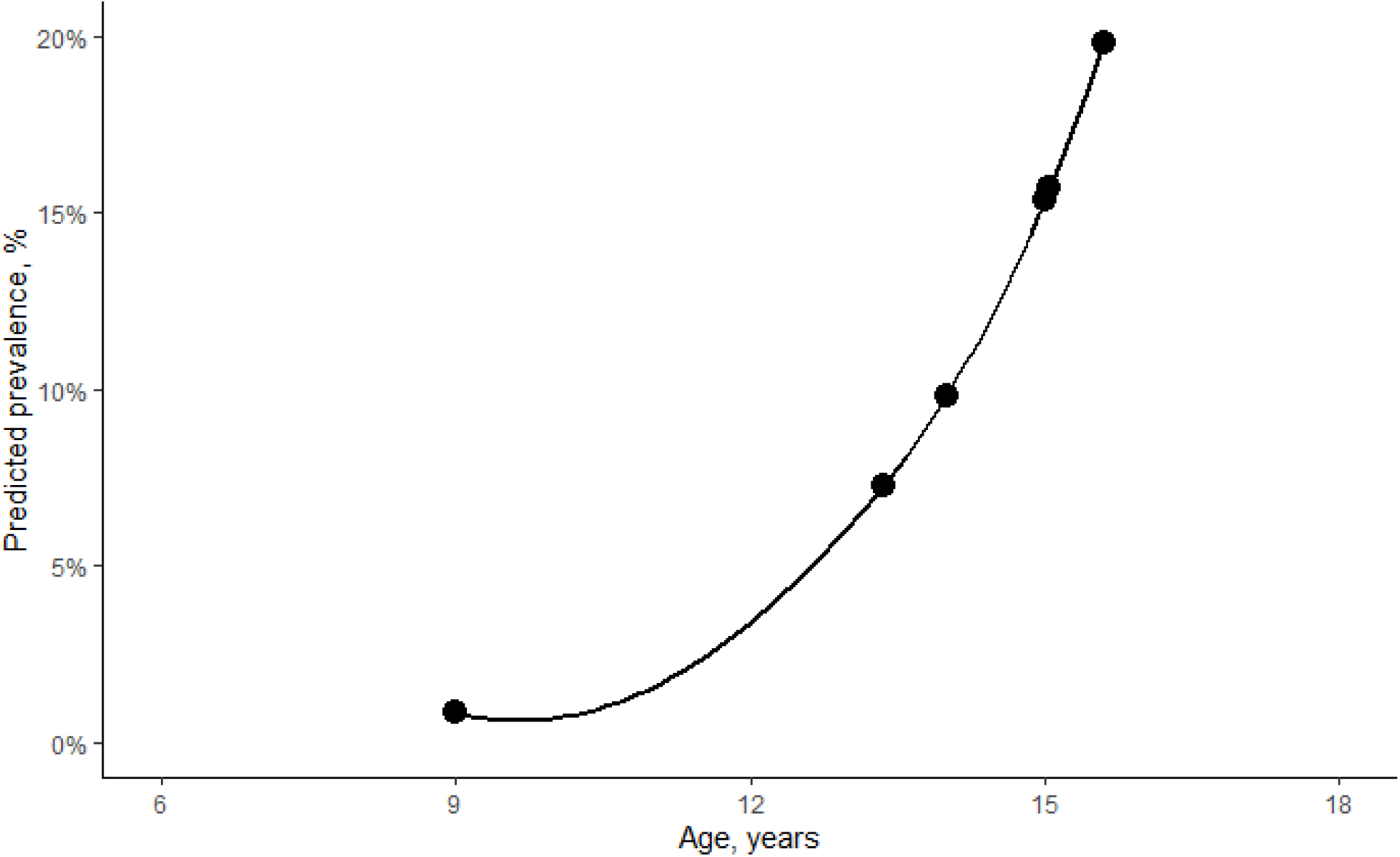
Relation of age and prevalence of depressive disorders

### Mood and anxiety disorders

Four studies assessed generalized anxiety disorder (GAD). These studies were conducted in primary schools^45^, Almajiri schools^45^, secondary schools^31,38^, and primary care clinics^46^. One study was conducted among two subgroups of young children of primary school age in Kaduna: children attending Almajiri schools (N=213) and those attending conventional primary schools (N=200). Based on this study, the prevalence of GAD was 8.0% (95% CI: 4.6, 12.7) among primary school children and 16.0% (95% CI: 11.3, 21.6) among Almajiri children. This study also assessed the prevalence of other mental illnesses among the primary and Almajiri school children. Of these, GAD (8.0% vs. 16.0%), and agoraphobia (0.5% vs. 1.0%) were more common among Almajiri children. Separation anxiety (14.0% vs. 1.4%) and social phobia (2.5% vs. 0.9%) were less common among Almajiri children.

One study reported the prevalence of GAD, based on the PHQ-9, to be 19.1% and 17.9% respectively among private and public secondary school students in Osun State ^35^. Two studies in Ebonyi and Anambra states reported the prevalence of GAD among secondary school students to be 2.0% (n=560) and 10.1% (n=1,157), respectively. GAD prevalence in the Ebonyi state study was based on the 12-item general health questionnaire ^31^, while the the GAD-7 questionnaire ^46^ was used in the Anambra study. A pooled prevalence estimate of 9.5% (95% CI: 3.9, 21.4; I^2^ = 97%; 2,195 people; low certainty evidence – downgraded for serious imprecision) was obtained through a meta-analysis of all three studies examining the prevalence of GAD in secondary schools.

One study assessed GAD among children 7 – 14 years old attending a primary care clinic in Kwara and reported a prevalence of 1.1% (95% CI: 0.3, 2.9) ^46^. The assessment was done using the Rutter scale A2 and the Kiddie Schedule for Affective Disorders and Schizophrenia, Present and Lifetime Version (K-SADS-PL.

Merging the data from settings most representative of the general population of children and adolescents since 2010 – primary and secondary schools, in the absence of community studies, the pooled prevalence of GAD was 9.2% (95% CI: 4.5, 17.9; I^2^ = 95.9%; five studies among 2,395 individuals). The certainty of the evidence was very low – downgraded for inconsistency and serious imprecision. There was no relationship between the prevalence of GAD and the mean age (*p-*heterogeneity = 0.44).

In sensitivity analysis, we included studies conducted after 2006^47–49^, the year the most recent national survey was conducted. The pooled prevalence of GAD did not change materially: 6.2% in secondary schools (95% CI: 3.2, 11.7; I^2^ = 97%; five studies^31,35,46–48^ among 4,170 people). We now obtained pooled prevalence estimates for SAD: 2.4% in secondary schools (95% CI: 1.8, 3.2; I^2^ = 0%; two studies^47,48^ among 1,975 people)

Taken together, we infer that the prevalence of GAD was higher in studies conducted in schools.

### Neurodevelopmental disorders

Three studies assessed neurodevelopmental disorders. These studies were conducted in primary schools ^28,45^ and in primary care settings ^38^.

The prevalence of ADHD was 2.8% among 200 primary school children in Zaria, Kaduna ^45^ and 2.7% among primary school children in Ikot-Ekpene, Akwa-Ibom^28^. ADHD was assessed using different instruments in both studies: the K-SADS-PL^45^ and the Vanderbilt ADHD Diagnostic Teacher Scale^28^. The pooled prevalence from both studies was 2.8% (95% CI: 2.0, 3.8; I^2^ = 0%; 1,374 people; high certainty evidence). In sensitivity analysis, including one study conducted in 2006^50^ changed the pooled prevalence meaningfully (5.8%; 95% CI: 2.9, 11.6; I^2^ = 97%; three studies among 3,959 people).

In the Zaria study, the prevalence of ADHD was 0.5% among Almajiri children^45^. The prevalence of ADHD in primary care settings was 1.4% (95% CI: 0.5, 3.3) based on a single study among 350 children 7 – 14 years of age in Kwara^38^. ADHD was assessed using the K-SADS-PL in this study.

Autism spectrum disorder among children and adolescents was examined in one study conducted among primary and secondary school students in Ebonyi and Enugu states ^51^. The prevalence of autism spectrum disorders among in this study^51^ was 2.9% (95% CI: 1.8, 4.4).

### Other mental disorders

Four studies assessed disruptive, impulse control and conduct disorders. Among 9,458 secondary school students across 47 schools in Lagos^29^, the prevalence of behavioral disorder was 15.1% (95% CI: 14.4, 15.8). Among 1,174 primary school children in Akwa Ibom^28^, the prevalence of conduct disorder was 9.8% (95% CI: 8.2, 11.6). In the Zaria study^45^, the prevalence of oppositional defiant disorder was 1.5% (95% CI: 0.3, 4.3) among 200 primary school children and 1.4% among 213 Almajiri children ^45^. In the same study, the prevalence of obsessive compulsive disorder (OCD) was found to be 2.5% among primary school children and 0% among Almajiri children^45^.

Among 147 juveniles remanded to a correctional facility in Ogun ^52^, the prevalence of conduct disorder was 56.5% (95% CI: 48.0, 64.6).

Post-traumatic stress disorder (PTSD) was assessed by five studies across different settings. The first study conducted among 560 adolescents and young adults aged 14 – 22 years attending secondary school in Ebonyi^31^ found a PTSD prevalence of 7.9% (95% CI: 5.8, 10.4). Another study evaluated PTSD among 144 juvenile offenders, 16 – 20 years old, in Abeokuta, Ogun State^53^, and found a prevalence of 9.7% (95% CI: 5.4, 15.8). One study assessed PTSD among 287 adolescents in an unspecified trauma-affected community^54^ and found a PTSD prevalence of 15.3% (95% CI: 11.4, 20.0). Another study assessed PTSD using the NIMH Diagnostic Interview Schedule for children among 73 adolescents at a camp for internally displaced persons in Kaduna and found a prevalence of 3% ^55^. The highest prevalence of PTSD was reported in a study among 449 out-of-school and unstably housed adolescents in Ibadan, Oyo^56^: 29.6% (95% CI: 25.4, 34.1).

Two studies assessed substance use disorders. These studies were conducted in secondary schools^29^, and correctional institutions^57^. The first study was among 9,458 students aged 11 – 21 y old across 47 public secondary schools in Lagos. The prevalence of substance use disorders in this study was 2.4% (95% CI: 2.1, 2.7; high certainty). This study also found the prevalence of alcohol use disorder to be 4.3% (95% CI: 3.9, 4.7). The second study was conducted among 165 male and female adolescents (14 – 17 y) remanded to correctional institutions in Lagos^58^. The prevalence of substance use disorder among adolescents in correctional institutions was found to be 15.8% (95% CI: 10.6, 22.2).

Two studies assessed schizophrenia spectrum disorders. These studies were conducted among primary^45^ and secondary school ^29^ children and adolescents. Among 413 children attending Almajiri model and conventional primary schools in Kaduna^45^, the prevalence of psychosis was 0% (95% CI: 0%, 1.8%) in conventional primary schools and was 2.3% (95% CI: 0.8, 5.4) among Almajiri children. Among 9,441 secondary school students 11 – 21 years old in Lagos, psychotic-like experiences were reported in 10.5% (95% CI: 9.9, 11.1), based on the MINI-Kid.

### Mental disorders by setting

**Figure 4** presents the best available evidence of the prevalence of mental disorder studied among children and adolescents in Nigeria since 2010. In primary care clinics **(Figure 4A)**, moderate certainty evidence indicates the prevalence was 1.4% for ADHD, 0.6% for mental retardation and 0.6% for autism spectrum disorders. In primary schools (**Figure 4B),** high certainty evidence indicates the prevalence was 0.4% for major depression and 2.8% for ADHD. Moderate certainty evidence indicates the prevalence was 8.0% for GAD, 14% for separation anxiety and 9.8% for conduct disorder. Among secondary school students (**Figure 4C)**, high certainty evidence indicates the prevalence was 2.4% for substance use disorder, 10.5% for psychotic-like experiences and 15.1% for behavioral disorder. Moderate certainty evidence indicates the prevalence was 7.9% for PTSD. Among children with chronic diseases (**Figure 4D),** the prevalence of major depression was 15.1% for HIV, based on high certainty evidence, and 30.8% for epilepsy, based on moderate certainty evidence.

**Figure 4.**
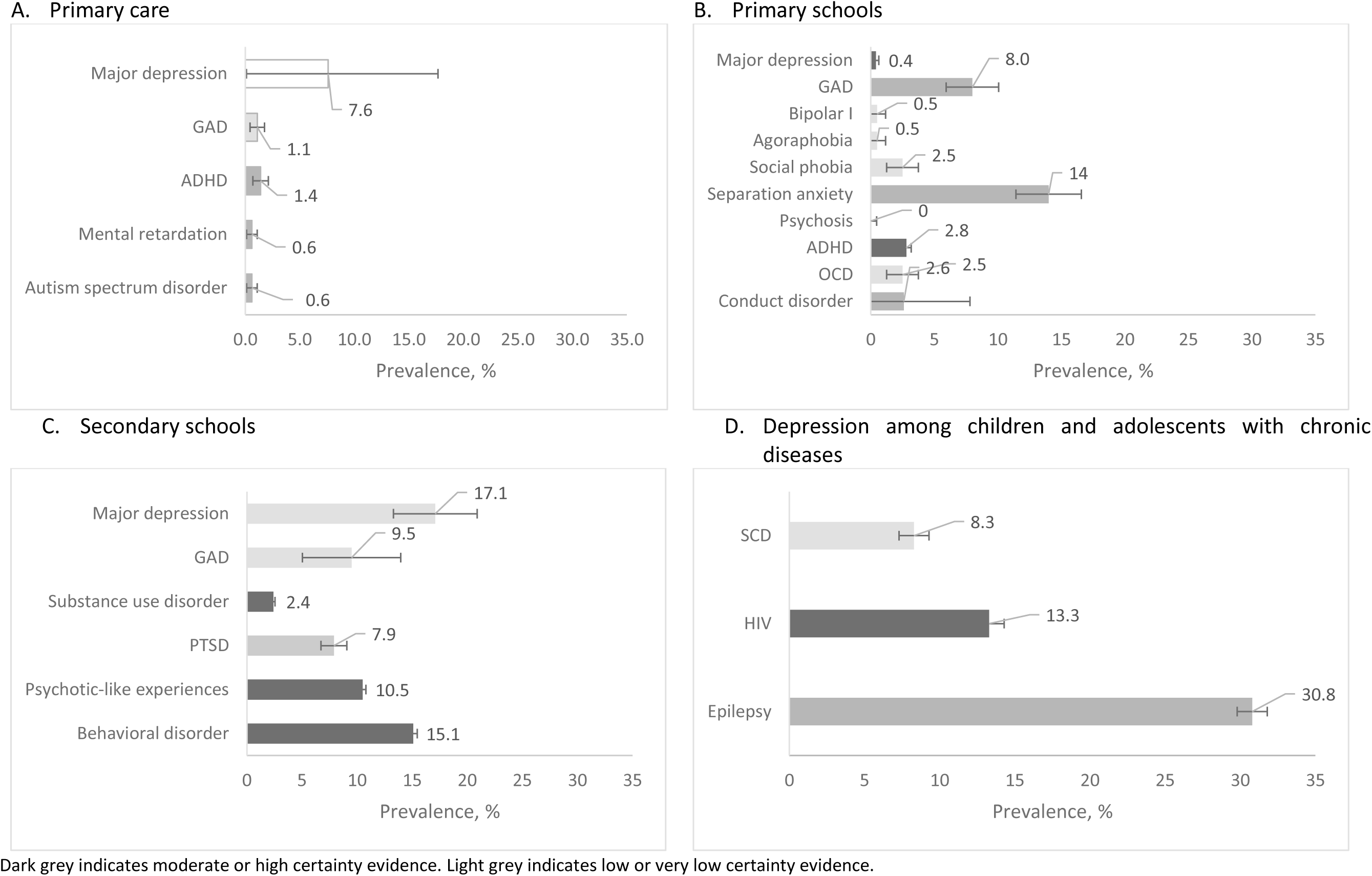
Mental disorder among children and adolescents in Nigeria based on the best available evidence

## Discussion

This systematic review and meta-analysis provides a comprehensive evaluation of the prevalence of mental health disorders among pediatric populations in Nigeria. Our findings highlight the burden of major depressive disorder (MDD), generalized anxiety disorder (GAD), attention-deficit/hyperactivity disorder (ADHD), and behavioral disorders among Nigerian children and adolescents. Study results reflect mental health challenges within Nigeria but demonstrate alignment with broader global patterns, underscoring the universal nature of mental health in pediatric populations.

We observed in our analyses that the prevalence of depression in Nigeria tends to be low in childhood, but increases sharply during adolescence. This is supported by findings in extant literature^59^. We found that about 1 in 5 secondary school students in Nigeria (17%) may suffer from depression. Adolescent depression significantly elevates the risk of negative outcomes, with studies indicating that depressed youths are more likely to experience academic failure^60^, have lower lifetime earnings^61^, and more likely to engage in substance abuse^62^, and attempt suicide^63^ compared to their non-depressed peers. Adolescent depression has also been shown to be a predictor of psychiatric disorders in adulthood ^60–62^.

Although the estimated prevalence of depression in Nigeria aligns with global prevalence patterns e.g., 12% in Uganda, and 11% in the USA^1^, childhood and adolescent depression in Nigeria is noted to be markedly higher than in some countries, particularly those in Europe. The prevalence of major depression among Nigerian secondary school students was found to be 17% in our study, but population studies and systematic reviews across Europe indicate that the prevalence in similar age groups may be as low as 1.7% ^66^. This disparity in prevalence may be due to differences in diagnostic practices, cultural perceptions of mental health, and increased access to mental health services, which allow for earlier detection, prompt treatment, and less severe disease in European countries ^67^. In Nigeria, mental health disorders are often underreported in healthcare facilities due to limited mental health literacy and concerns about stigma ^68,69^. Addressing cultural barriers through targeted public health campaigns and education initiatives, and implementing interventions that improve access and quality of care would be essential for improving diagnosis and management of depression among Nigerian children and adolescents.

Studies on mood and anxiety disorders in children and adolescents in Nigeria are scarce. Hence prevalence estimates of generalized anxiety disorder, separation anxiety disorder and social phobia, in this study were based on single study estimates. The impact of these disorders on life outcomes, including quality of life is known to be significant. Childhood anxiety predicts the occurrence of mental health disorders in adolescence, including depression^70^. Multiple factors contribute to childhood anxiety disorders including environmental stress, parenting styles and changes in the brain amygdala and subcortical structures during maturation^71^. More studies may be needed to establish the prevalence of these disorders in the Nigerian pediatric population. Policies and programs aimed at preventing and treating anxiety disorders in children and adolescents should also be given adequate prioritization.

There has been an explosion in global interest in ADHD over the last decade, with tens of thousands of research articles published on the subject in that time^72^. Unfortunately, we observed that only few relevant studies have been conducted in Nigeria since 2010. Based on two studies among primary school children, we estimate that the prevalence of ADHD among children in Nigeria is approximately 2.8% – lower than global estimates of 5–8% ^72,73^. A low diagnosed prevalence may reflect the actual burden of disease or may be due to low access of trained personnel or cultural factors that influence the perception and reporting of ADHD symptoms ^74^. ADHD is associated with severe consequences for affected individuals including learning disabilities, education attainment gaps, and higher costs of education and healthcare^75^. Untreated ADHD can lead to significant long-term impairments in educational and social functioning, and a higher risk of substance abuse, and involvement in criminal activities later in life^76^. These findings highlight the urgent need for improved awareness and diagnostic practices for ADHD in Nigeria and other LMICs.

We found behavioral disorders to be the most common mental disorders among adolescents in Nigeria, with a best-available prevalence of 15.1%. This finding aligns with data from a meta-analysis of studies from multiple countries which found a similar prevalence ranging from 10% to 16% ^77^. The high prevalence of behavioral disorders in Nigerian adolescents is particularly concerning, given the strong association between these disorders and adverse medical and humanistic outcomes ^78^. The epidemiology of behavioral disorders may be influenced by socio-economic factors, including poverty, greater exposure to violence, and limited access to mental health care—factors that are more prevalent in LMICs^79,80^. Behavioral disorders in adolescence are often associated with broader social and economic costs, as affected individuals are more likely to drop out of school, become involved in the criminal justice system, and face difficulties in employment ^81^. These further highlight the critical importance of early intervention and prevention strategies aimed at reducing the prevalence of mental disorders among young Nigerians.

Our analysis has noteworthy strengths and limitations. First, we carefully pooled studies by mental disorder and by setting, allowing us to make correct inferences, with limited assumptions about how findings from one setting may be generalizable to other settings. Second, we evaluated the certainty of the evidence using GRADE principles, and identified the best available evidence for each prevalence estimate from our analysis. Our study was limited by the availability and quality of primary studies for synthesis. Across many settings and disorders, there were either no studies or single studies, highlighting critical gap for future research. In addition, the included studies were not geographically representative of Nigerian states, and included studies were frequently conducted in urban settlements. By restricting our analysis to 2010, we are able to focus on current prevalence estimates, relevant for planning and policy today, but unable to examine the historical prevalence and trends of mental illness among children and adolescents in Nigeria. We were unable to examine publication bias due to the state of the methodology of prevalence meta-analyses, and considerable publication bias could meaningfully shift the pooled prevalence estimates from our studies in either direction. Our results highlight significant research gaps, particularly the urgent need for high-quality primary studies to inform policy and practice effectively.

## Conclusion

Our comprehensive assessment of mental disorders in Nigeria since 2010 reveals notably high prevalence rates for major depressive disorder, GAD, ADHD, and behavioral disorders, underscoring the substantial impact these conditions have on Nigerian children and adolescents. The prevalence of mental disorders varied substantially by the setting studied. Clinical and public health campaigns focused on early intervention and preventive strategies and tailored to appropriate settings may be necessary to address the root causes and reduce the incidence of mental disorders in Nigeria.

## Data Availability

All raw data used in this work are available upon reasonable request to the authors. However, all the results produced are all contained in the manuscript.

## Appendix tables

**Appendix 1.**
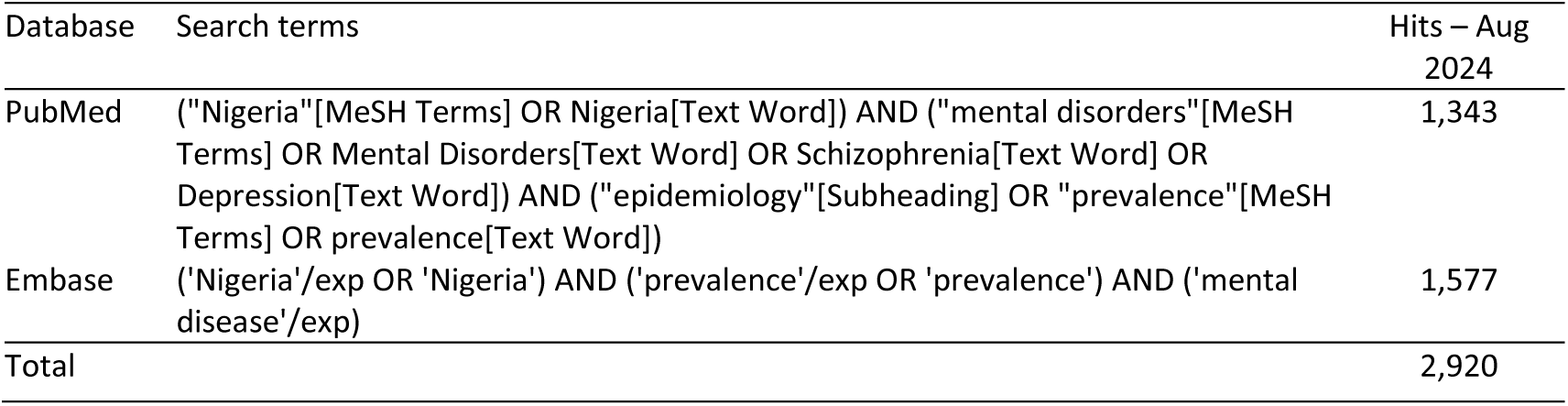
Search strategy.

**Appendix 2.** Instruments used to diagnose and categorize depression.

| Diagnosis | Instrument | Characteristics | Any depression | Moderate category | Severe category |
| --- | --- | --- | --- | --- | --- |
| Depression | Beck Depression Inventory <sup>83</sup> |  |  |  |  |
|  | BDI |  | >14 | >18 | >29 |
|  | BDI-II |  |  | >19 | >28 |
|  | Center for Epidemiologic Studies – Depression Scale (CES-D) <sup>84</sup> |  | >11 | >15 | >25 |
|  | Children’s Depression Inventory <sup>85</sup> |  |  | >20 |  |
|  | Diagnostic Interview Schedule for Children (DISC) |  |  | Clinician-rated |  |
|  | Goldberg Depression Questionnaire |  |  |  |  |
|  | Hopkins symptom checklist-25 (HSCL-D) |  | >1.75 |  |  |
|  | Hospital Anxiety and Depression Scale (HADS) <sup>86–88</sup> |  | ≥8 | >11 | ≥15 |
|  | Mini International Neuropsychiatric Interview (MINI) |  | Clinician-rated | Clinician-rated |  |
|  | Patient Health Questionnaire (PHQ-9) <sup>83</sup> |  | >5 | >10 | >20 |
|  | Schedule for Clinical Assessment in Psychiatry (SCAN) |  |  | Clinician-rated |  |
|  | Schedule for Affective Disorders and Schizophrenia for School-aged Children Present and Life- time Version (K-SADS-PL) |  |  | Clinician-rated |  |

**Appendix 3.** Community studies.

| Author, year | City | State | Study Design | Setting | Sample size | Age, mean, y; | % Female | Diagnosis | Description of assessment | Findings | Risk of bias |
| --- | --- | --- | --- | --- | --- | --- | --- | --- | --- | --- | --- |
| Ayamolowo, 2019 <sup>39</sup> | Ile-Ife | Osun | multistage sampling | Child-rearing teenagers in community | 120 |  | 100 | Borderline clinical depression | Beck's Depression Inventory | 11.67% | Fair |
|  |  |  |  |  |  |  |  | Moderate depression |  | 20.00% |  |
|  |  |  |  |  |  |  |  | Severe depression |  | 10.00% |  |
| Cheng, 2014 <sup>56</sup> | Ibadan | Oyo | Respondent-driven sampling (RDS) | out-of-school youth and unstably housed youth | 449 | 16.65 | 51.00% | PTSD | Post-Traumatic Stress Disorder (PTSD) Checklist-Civilian version (PCL-C, 17 items) | 29.62% | Fair |
| Sheikh, 2016 <sup>89</sup> | Kaduna | Kaduna | Total sampling | Internally displaced persons' (IDP) camp | 73 | 13 | 49.30% | Probable PTSD | Reaction of Adolescents to Traumatic Stress (RATS) instrument | 34.25% | Fair |
|  |  |  |  |  |  |  |  | Definitive PTSD | NIMH Diagnostic interview schedule for children | 2.74% |  |
| Stupar, 2021 <sup>54</sup> | Nigeria |  | simple random | rural and urban communities | 287 | 14.54 | 58.5 | Post-traumatic stress disorder | UCLA PTSD Reaction Index | 15.33% | Good |

**Appendix 4.**
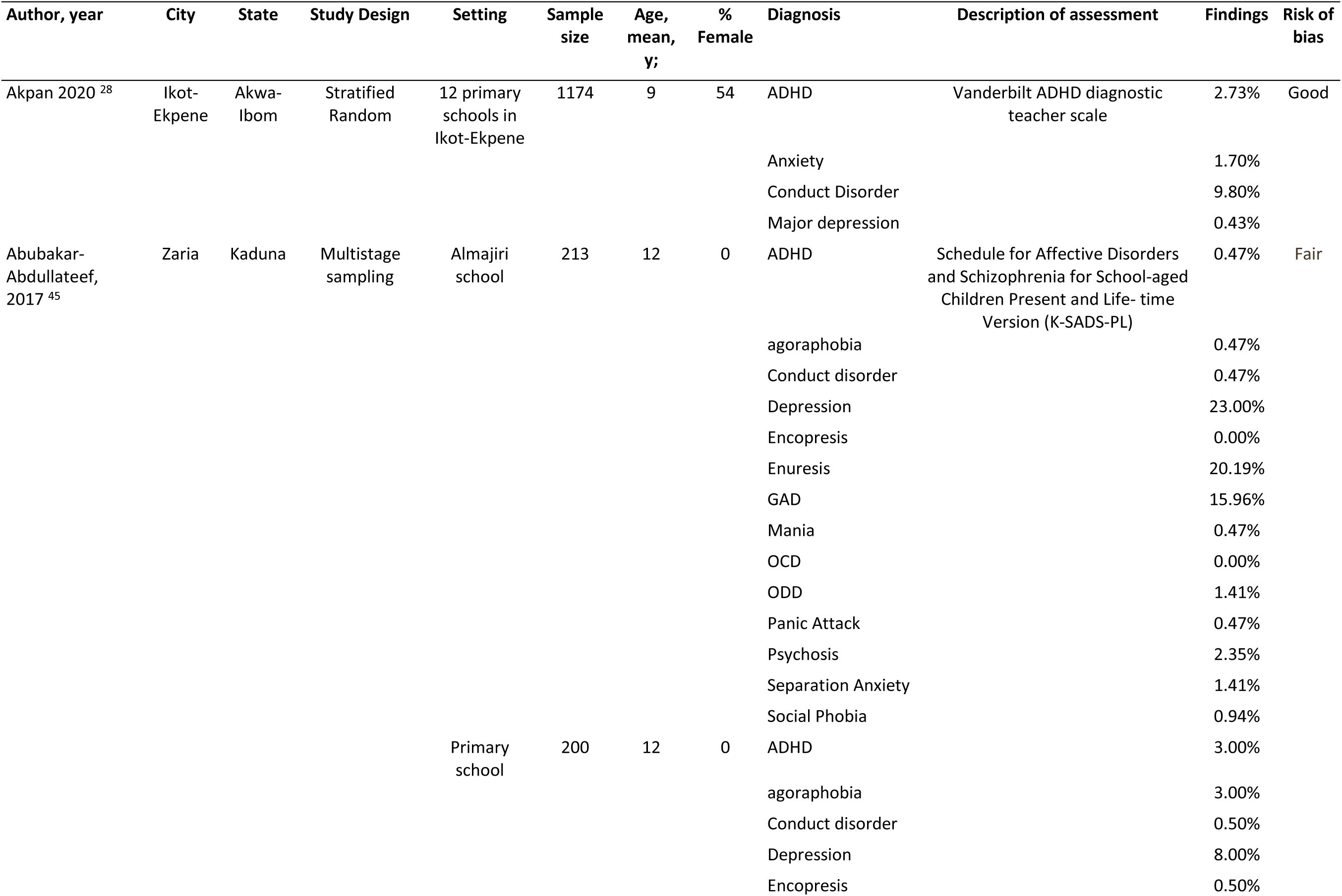
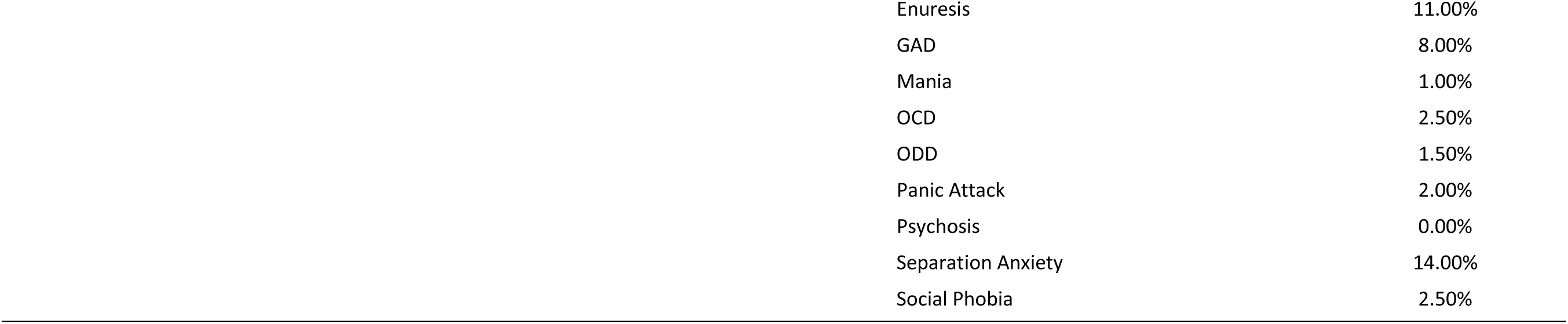
Primary school children – included studies.

| Author, year | City | State | Study Design | Setting | Sample size | Age, mean, y; | % Female | Diagnosis | Description of assessment | Findings | Risk of bias |
| --- | --- | --- | --- | --- | --- | --- | --- | --- | --- | --- | --- |
| Akpan 2020 <sup>28</sup> | Ikot-Ekpene | Akwa-Ibom | Stratified Random | 12 primary schools in Ikot-Ekpene | 1174 | 9 | 54 | ADHD | Vanderbilt ADHD diagnostic teacher scale | 2.73% | Good |
|  |  |  |  |  |  |  |  | Anxiety |  | 1.70% |  |
|  |  |  |  |  |  |  |  | Conduct Disorder |  | 9.80% |  |
|  |  |  |  |  |  |  |  | Major depression |  | 0.43% |  |
| Abubakar-Abdullateef, 2017 <sup>45</sup> | Zaria | Kaduna | Multistage sampling | Almajiri school | 213 | 12 | 0 | ADHD | Schedule for Affective Disorders and Schizophrenia for School-aged Children Present and Life- time Version (K-SADS-PL) | 0.47% | Fair |
|  |  |  |  |  |  |  |  | agoraphobia |  | 0.47% |  |
|  |  |  |  |  |  |  |  | Conduct disorder |  | 0.47% |  |
|  |  |  |  |  |  |  |  | Depression |  | 23.00% |  |
|  |  |  |  |  |  |  |  | Encopresis |  | 0.00% |  |
|  |  |  |  |  |  |  |  | Enuresis |  | 20.19% |  |
|  |  |  |  |  |  |  |  | GAD |  | 15.96% |  |
|  |  |  |  |  |  |  |  | Mania |  | 0.47% |  |
|  |  |  |  |  |  |  |  | OCD |  | 0.00% |  |
|  |  |  |  |  |  |  |  | ODD |  | 1.41% |  |
|  |  |  |  |  |  |  |  | Panic Attack |  | 0.47% |  |
|  |  |  |  |  |  |  |  | Psychosis |  | 2.35% |  |
|  |  |  |  |  |  |  |  | Separation Anxiety |  | 1.41% |  |
|  |  |  |  |  |  |  |  | Social Phobia |  | 0.94% |  |
|  |  |  |  | Primary school | 200 | 12 | 0 | ADHD |  | 3.00% |  |
|  |  |  |  |  |  |  |  | agoraphobia |  | 3.00% |  |
|  |  |  |  |  |  |  |  | Conduct disorder |  | 0.50% |  |
|  |  |  |  |  |  |  |  | Depression |  | 8.00% |  |
|  |  |  |  |  |  |  |  | Encopresis |  | 0.50% |  |

|  |  |
| --- | --- |
| Enuresis | 11.00% |
| GAD | 8.00% |
| Mania | 1.00% |
| OCD | 2.50% |
| ODD | 1.50% |
| Panic Attack | 2.00% |
| Psychosis | 0.00% |
| Separation Anxiety | 14.00% |
| Social Phobia | 2.50% |

**Appendix 5.**
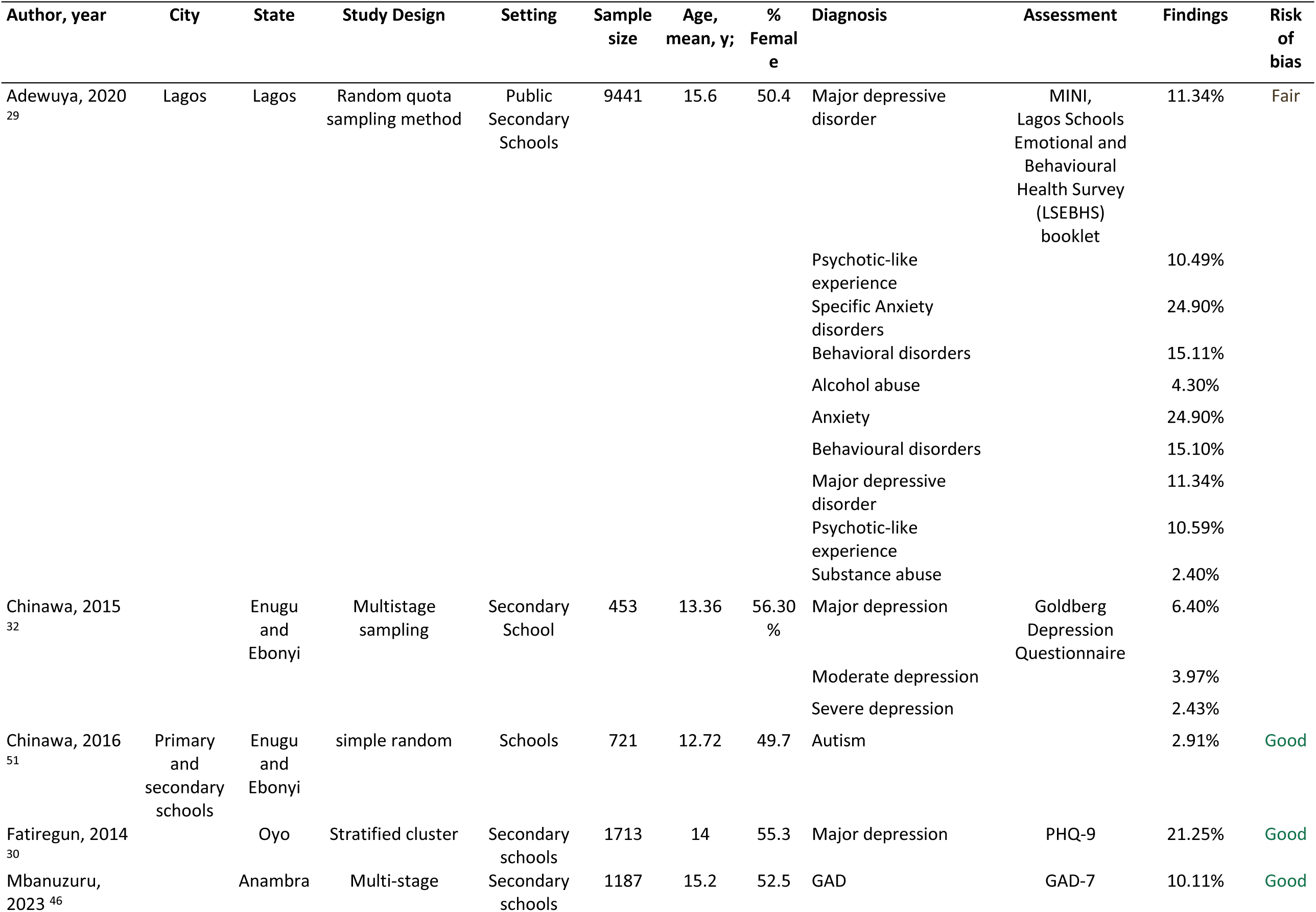
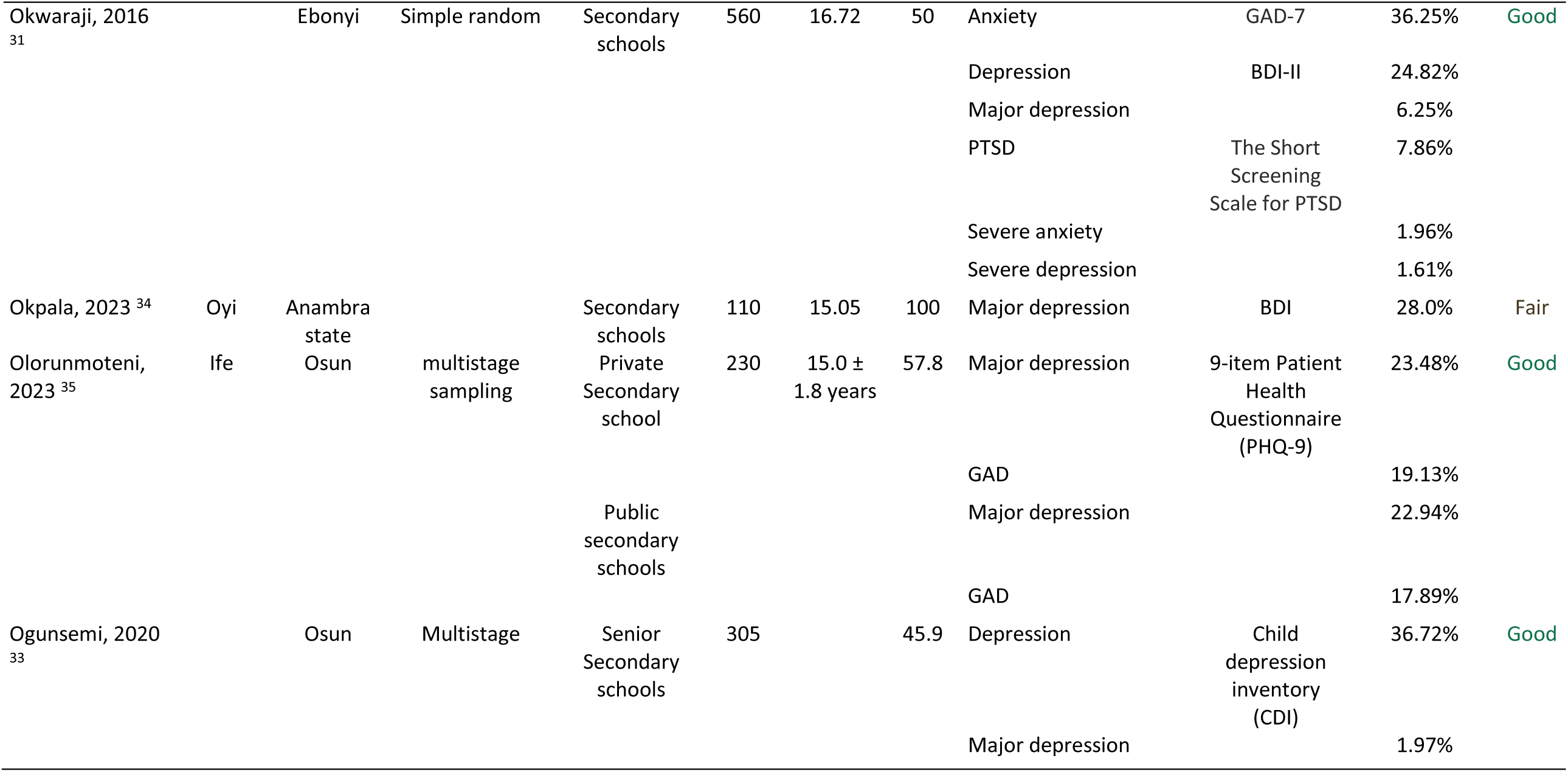
Secondary school students – included studies.

**Appendix 6.** Primary care – Included studies.

| Author, year | City | State | Study Design | Setting | Sample size | Age, mean, y; | % Female | Diagnosis | Description of assessment | Findings | Risk of bias |
| --- | --- | --- | --- | --- | --- | --- | --- | --- | --- | --- | --- |
| Oladeji, 2019<sup>36</sup> | Ibadan | Oyo | Random | Teen moms | 772 | 17.8 | 100% | Major depression | Edinburgh Postnatal Depression Scale (EPDS) | 17.75% | Fair |
| Abiodun 2011<sup>38</sup> | Ilorin | Kwara | Total | Primary care setting | 350 | 9.75 | 48.30% | ADHD | K-SADS-PL | 1.43% |  |
|  |  |  |  |  |  |  |  | GAD |  | 1.14% |  |
|  |  |  |  |  |  |  |  | Conduct disorder |  | 1.71% |  |
|  |  |  |  |  |  |  |  | Major depression |  | 0.57% |  |
|  |  |  |  |  |  |  |  | Enuresis |  | 6.00% |  |
|  |  |  |  |  |  |  |  | Mental retardation |  | 0.57% |  |

**Appendix 7.**
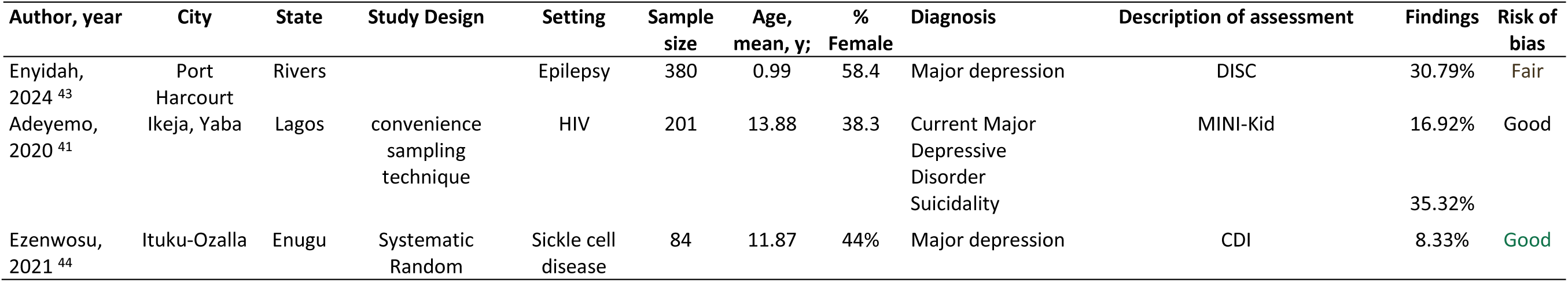
Children with chronic illnesses – included studies.

**Appendix 8.**
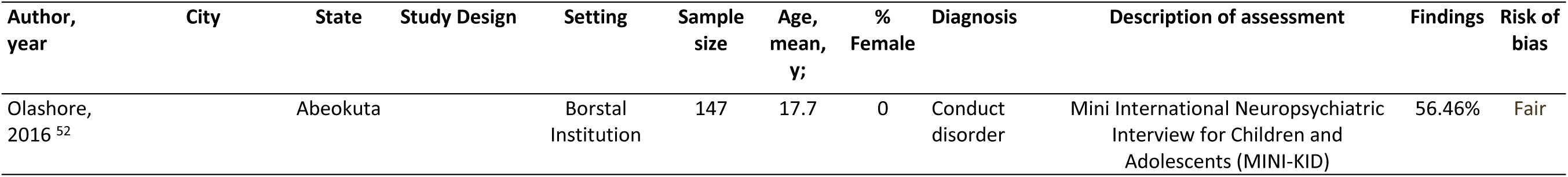
Correctional facilities – included studies.

| Author,<br>year | City | State | Study Design | Setting | Sample<br>size | Age,<br>mean,<br>y; | %<br>Female | Diagnosis | Description of assessment | Findings | Risk of<br>bias |
| --- | --- | --- | --- | --- | --- | --- | --- | --- | --- | --- | --- |
| Olashore,<br>2016 <sup>52</sup> |  | Abeokuta |  | Borstal<br>Institution | 147 | 17.7 | 0 | Conduct<br>disorder | Mini International Neuropsychiatric<br>Interview for Children and<br>Adolescents (MINI-KID) | 56.46% | Fair |

**Supplementary figure 1:**
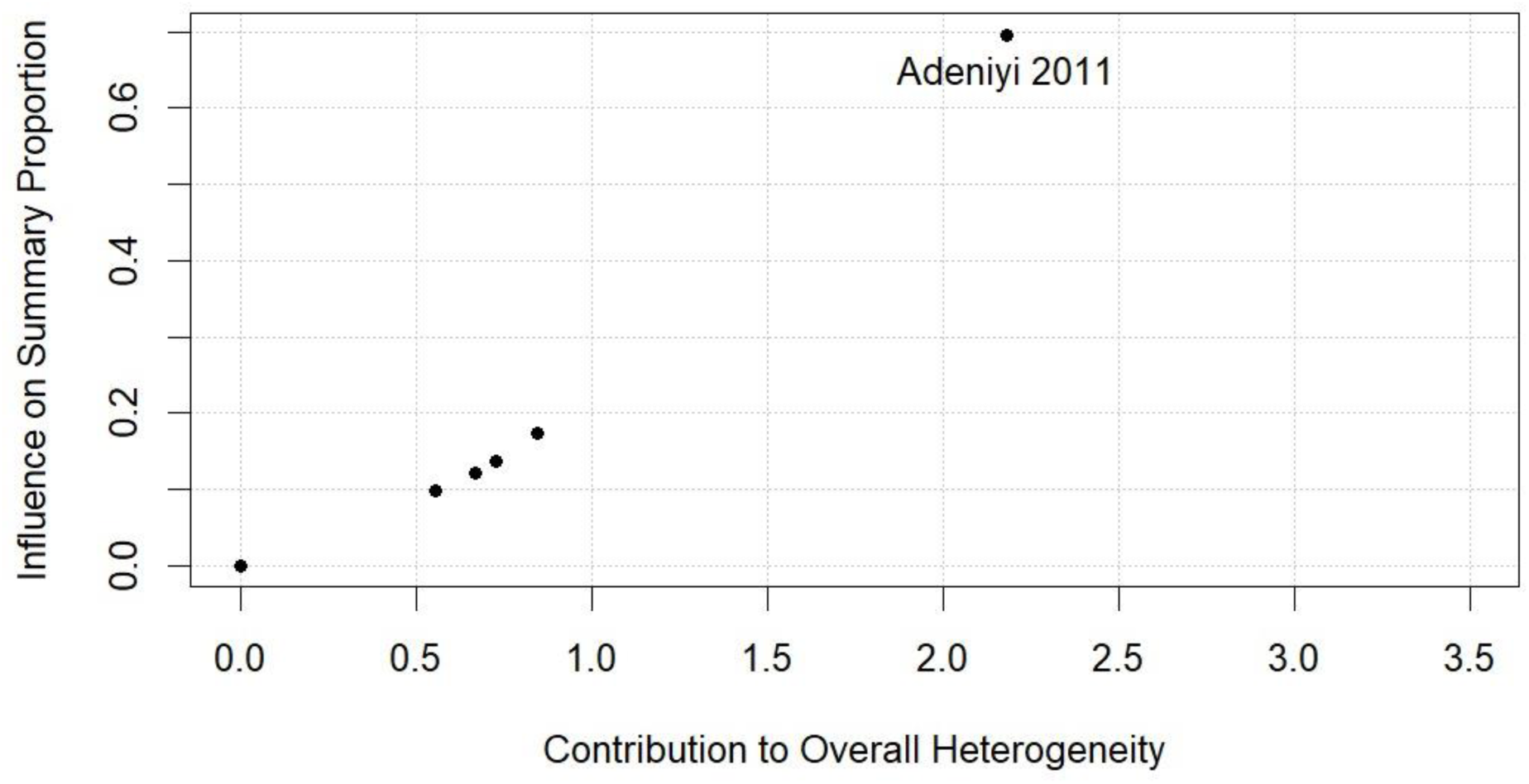
relationship between the influence of the individual studies on the summary proportion against their contribution to the overall heterogeneity

